# Development of a Prognostic Model for Sepsis Based on Gut Microbiota-Associated Genes and Identification of Potential Targets

**DOI:** 10.1101/2025.11.17.25340434

**Authors:** Fangqiong Li, Minrong Xu, Huiqin Xiao, Ping Hu, Wei Zhang

## Abstract

**Background:** Gut microbiota dysbiosis drives sepsis progression by impairing intestinal barrier function and exacerbating systemic inflammation, but the microbiota-host-immune interaction mechanisms remain unclear.

**Methods:** We integrated transcriptomic and single-cell RNA sequencing (scRNA-seq) data from the Gene Expression Omnibus (GEO) database. Differentially expressed genes (DEGs) between sepsis patients and healthy controls were identified in GSE154918, then intersected with 248 gut microbiota-related genes from the GutMGene database to obtain candidate genes. A prognostic model named GMGscore was constructed via LASSO-Cox regression in GSE65682 and validated in GSE95233. Area under the curve (AUC) was used to evaluate the model performance. scRNA-seq data (GSE167363) was used to determine the cellular localization of key genes. Molecular docking predicted interactions between gut microbiota metabolites and the key target.

**Results:** We identified 34 gut microbiota-related DEGs, which were enriched in pathways like inflammatory bowel disease and IL-17 signaling. The GMGscore, based on 6 genes (CYP1A2, FFAR2, IL4R, MUC1, RORA, ASPM), showed excellent prognostic performance (AUC = 0.903 in training set; AUC = 0.901 in validation set). High GMGscore correlated with poor survival, upregulated neutrophil degranulation and reduced neutrophils. RORA was identified as a key gut microbiota-related target, which was consistently downregulated in sepsis with the highest diagnostic AUC across datasets, mainly expressed in effector T cells and NK cells, and positively correlated with CD8+ T cell/NK cell infiltration (R = 0.419 and 0.352, respectively). Virtual knockout of RORA downregulated cytotoxic genes (CCL5, NKG7, GNLY). Molecular docking showed stable binding of RORA with *Collinsella*-derived metabolites (Citric acid, Sedoheptulose, and Tricarballylic acid).

**Conclusions:** The GMGscore is a robust prognostic tool for sepsis. RORA, targeted by gut microbiota metabolites, may regulate immune balance via effector T cells and NK cells. These findings advance understanding of gut microbiota-sepsis crosstalk and provide new avenues for precise prognosis and targeted therapy.

## Introduction

Sepsis is one of the leading causes of death in intensive care units (ICUs) worldwide, characterized by an excessive inflammatory response and immune suppression imbalance triggered by infection (1). According to epidemiological reports, sepsis has been recognized as a global health burden since 2017 due to its high incidence and mortality rates, with an estimated global incidence of approximately 49 million cases and a mortality rate as high as 20%-30% (2). The annual healthcare costs associated with sepsis are estimated to be $24 billion (3, 4). Although the Surviving Sepsis Campaign (SSC) guidelines recommend early anti-infective therapy and fluid resuscitation (5), sepsis patients exhibit significant individual variability. Conventional prognostic markers, such as procalcitonin and C-reactive protein, lack sufficient specificity, making it difficult to achieve accurate risk stratification (6). Therefore, the development of prognostic models and therapeutic strategies based on molecular mechanisms is a critical need for reducing the mortality rate of sepsis (7, 8).

Dysbiosis of the gut microbiota is one of the core drivers of the pathological progression of sepsis. Under septic conditions, impairment of the intestinal barrier function leads to bacterial translocation and the entry of harmful metabolites into the bloodstream, which exacerbates the systemic inflammatory response and forms a vicious cycle (8–11). Previous studies have confirmed that the composition and metabolites of the gut microbiota may affect an individual’s immune phenotype and the prognosis of sepsis (12–14). Thus, the development of gut microbiota-associated biomarkers can provide a more comprehensive perspective for the prognostic evaluation of sepsis (15). In addition, gut microbiota-related interventions have been shown to improve intestinal barrier function and survival rates in septic mice (12, 16, 17). However, the interaction mechanisms among the gut microbiota, host genes, and immune system in sepsis, as well as the key regulatory targets, remain unclear, which limits the clinical translation of precise interventions.

In this study, through multi-omics integrated analysis, we aim to find out the core prognostic genes associated with the gut microbiota in sepsis, construct a quantifiable prognostic score, designated as GMGscore, and identify potential targets that can be intervened via gut microbiota modulation and explore their underlying molecular mechanisms.

## Materials and Methods

The complete flow chart of this study is shown in Fig. 1.

**Fig. 1.**
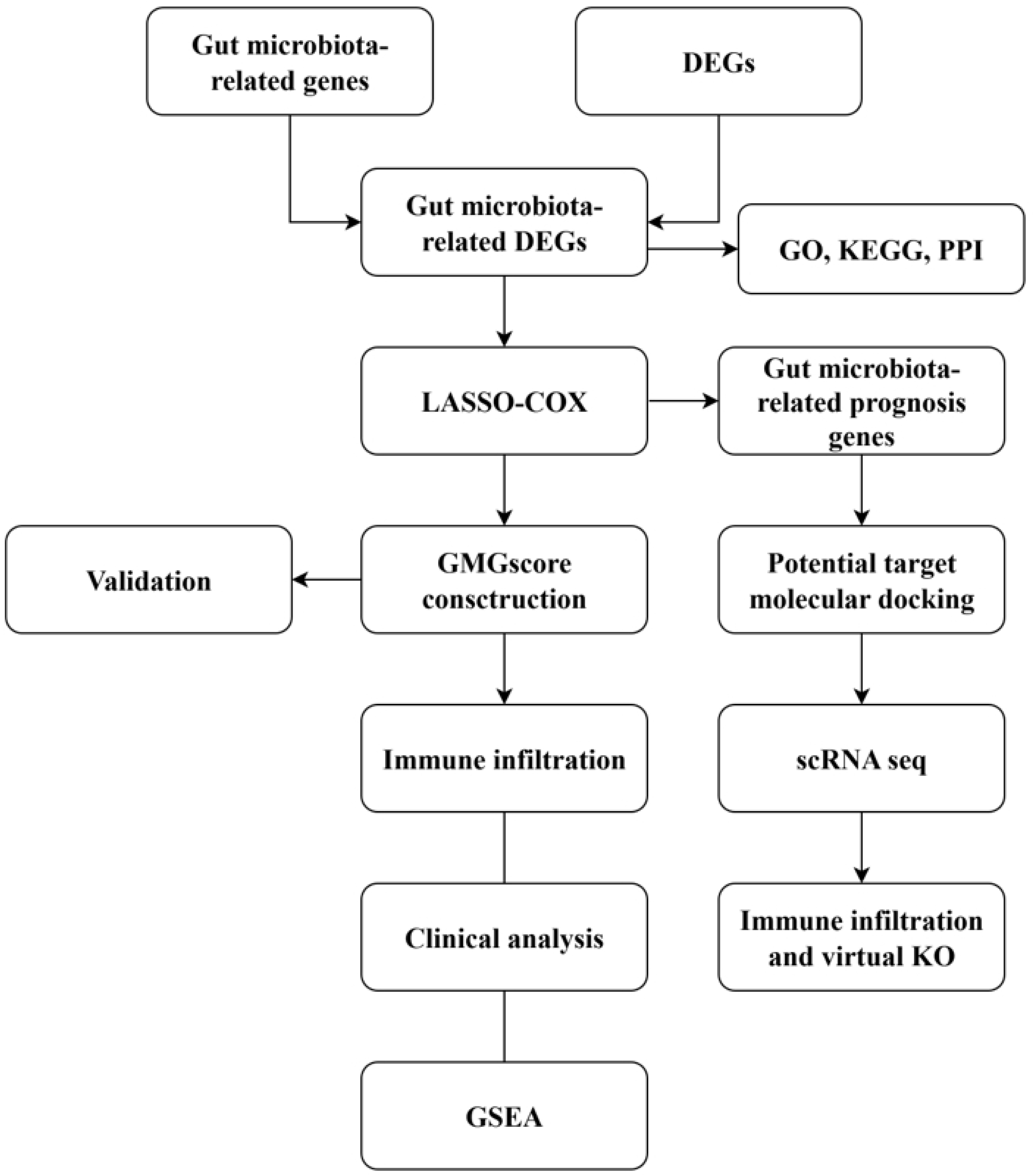
Flow chart of this current study.

### Data Sources

All datasets used in this study were retrieved from the Gene Expression Omnibus (GEO) database (https://www.ncbi.nlm.nih.gov/geo/). Only peripheral blood samples (whole blood or peripheral blood mononuclear cells) were included. The detailed information of all datasets is provided in Table S1.

The GutMGene database (http://bio-computing.hrbmu.edu.cn/gutmgene/#/home) provides extensive research resources on the interactions between the human gut microbiota and genes (18). A total of 248 genes closely associated with the gut microbiota and its metabolites were obtained from this database, with detailed information available in Table S2.

### Data Preprocessing

Data normalization and quality control were performed using R software (Version 4.3.1): Low-quality cells were filtered out using the “Seurat” package (19) with the following criteria: mitochondrial gene content >10% and unique molecular identifier (UMI) count <200. The “NormalizeData” and “FindVariableFeatures” functions in Seurat were applied to normalize the count data and identify highly variable genes, respectively. Cell clusters were visualized via Uniform Manifold Approximation and Projection (UMAP) with a resolution of 1.0.

Marker genes were detected using the “FindAllMarkers” function in Seurat based on the Wilcoxon rank-sum test. Only markers associated with clusters that met the threshold of log2 fold change > 0.25 were retained. After clustering, cells were reclassified into subpopulations, and the identity of each subpopulation was determined based on the similarity of gene expression patterns. Cell type annotation was conducted by combining the “SingleR” package (20) and the CellMarker database (http://biocc.hrbmu.edu.cn/CellMarker/).

### Screening of Candidate Genes

Differentially expressed genes (DEGs) between sepsis patients and healthy controls in GSE154928 was analyzed using the “DESeq2” package (21). DEGs were screened under the criteria of |log2 fold change| > 1.5 and adjusted P-value < 0.05. Candidate genes associated with the gut microbiota were obtained by taking the intersection of “DEGs” and “gut microbiota-associated genes”.

### Functional and Pathway Enrichment Analyses, and Construction of Protein-Protein Interaction (PPI) Network

Gene Ontology (GO) analysis is a commonly used method to investigate gene expression based on cellular functions or localization, typically conducted at three levels: Biological Process (BP), Molecular Function (MF), and Cellular Component (CC). The Kyoto Encyclopedia of Genes and Genomes (KEGG) is a database that stores information on genomes, biological pathways, diseases, and drugs. ClusterProfiler is a bioinformatics tool that integrates multiple functional analysis methods, featuring high efficiency in enrichment analysis and providing effective result visualization. The R package “clusterProfiler” was used for GO function enrichment analysis and KEGG pathway enrichment analysis (22). Statistical significance was set at adjusted P < 0.05. The PPI network was constructed using the STRING database (https://string-db.org/) with a confidence score > 0.4.

### Construction and Validation of the Prognostic Model (GMGscore)

Using GSE65682 as the training set, LASSO (Least Absolute Shrinkage and Selection Operator)-Cox regression analysis was performed with the “glmnet” package (23). The optimal λ value was determined via 10-fold cross-validation to screen genes significantly associated with 28-day mortality in sepsis. The GMGscore for each patient was calculated based on the regression coefficients using the formula: GMGscore = ∑ (Exp_ᵢ_ × Coef_ᵢ_), where Coef_ᵢ_ represents the coefficient of the gene, and Exp_ᵢ_ denotes the standardized expression level of that gene.

The “pROC” package (24) was used to generate receiver operating characteristic (ROC) curves, and the area under the curve (AUC) was calculated to evaluate the predictive performance of GMGscore for 28-day mortality in sepsis. Patients were divided into high-risk and low-risk groups according to the median GMGscore. Kaplan-Meier (KM) curves were plotted using the “survival” package, and the log-rank test was used to compare differences in survival rates between the two groups.

A nomogram was constructed by integrating GMGscore with clinical variables. The clinical utility of the nomogram was validated through ROC analysis, calibration curves, and decision curve analysis (DCA).

### Gene Set Enrichment Analysis (GSEA) and Immune Infiltration Analysis

GSEA was used to identify significantly altered key biological processes and pathways in the high-GMGscore group. The CIBERSORT algorithm (25) was applied to quantify the relative infiltration abundance of 22 immune cell types in the training set. Spearman correlation analysis was performed to explore the associations between key genes and immune cells.

### Screening of Key Targets and Molecular Docking

Based on information on key genes from the GutMGene database, Cytoscape software was used to construct a “gut microbiota-metabolite-host gene” interaction network. Targets meeting all the following criteria were screened: Significantly associated with sepsis prognosis (P < 0.05); Consistently differentially expressed in sepsis samples across all datasets; The regulatory direction of the gene by gut microbiota/metabolites is consistent with the direction of change that improves prognosis.

Structures of targets protein and gut microbiota metabolites were obtained from the UniProt database (https://www.uniprot.org/) and PubChem website (https://pubchem.ncbi.nlm.nih.gov/). Molecular docking was performed using AutoDock Vina (Version 1.2.0), with a binding energy < −5 kcal/mol as the screening criterion. The binding mode was visualized using PyMOL. AutoDock (https://ccsb.scripps.edu/mgltools/downloads/) and PyMOL software were used to simulate the binding between target proteins and small-molecule drugs.

### Single-Cell RNA Sequencing (scRNA-seq) Data Analysis

Based on the scRNA-seq data from GSE167363, the expression distribution of key targets across different cell types was analyzed. Secondary dimensionality reduction and clustering were performed on cell types with high target expression to clarify the specific cell subpopulation localization of the targets.

The scTenifoldKnk method (26) was used to simulate target gene knockout and infer expression changes of downstream related genes. This method has been proven to be an efficient systematic tool for studying gene function, particularly suitable for research environments where real knockout (KO) experiments are not feasible. The single-cell dataset was normalized and preprocessed, and the scTenifoldKnk method was used to construct a gene regulatory network (GRN) that captures the regulatory relationships between genes in sepsis patients. In the constructed GRN, unsupervised virtual knockdown was performed by approximating the deletion of target gene nodes to identify genes with significantly altered regulatory relationships due to target gene knockdown. The significance of these genes was ranked based on Z-scores and P-values, and the expression change patterns of upregulated and downregulated genes were visualized.

### Statistical Analysis

All statistical analyses were performed using R software (Version 4.3.1). For the comparison of continuous variables between two groups, an independent samples t-test or the Wilcoxon rank-sum test was used to analyze differences of variables. In Spearman correlation analysis, a correlation coefficient (R) > 0.3 was considered the correlation threshold. All statistical P-values were two-sided, and P < 0.05 was considered statistically significant.

## Results

### Screening and Functional Characteristics of Candidate Genes

In the GSE154918 dataset, a total of 2,958 DEGs were identified between sepsis patients and healthy controls, including 1,705 upregulated genes and 1,253 downregulated genes (Fig. 2A). A heatmap was used to visualize the top 10 upregulated and downregulated genes ranked by fold change (Fig. 2B). By taking the intersection of these DEGs with 238 gut microbiota-associated genes, 34 gut microbiota-related DEGs were obtained as candidate genes (Fig. 2C). A PPI network diagram was constructed to illustrate the associations among these candidate genes (Fig. 2D), and the chromosomal localization of these genes was visualized (Fig. S1).

**Fig. 2.**
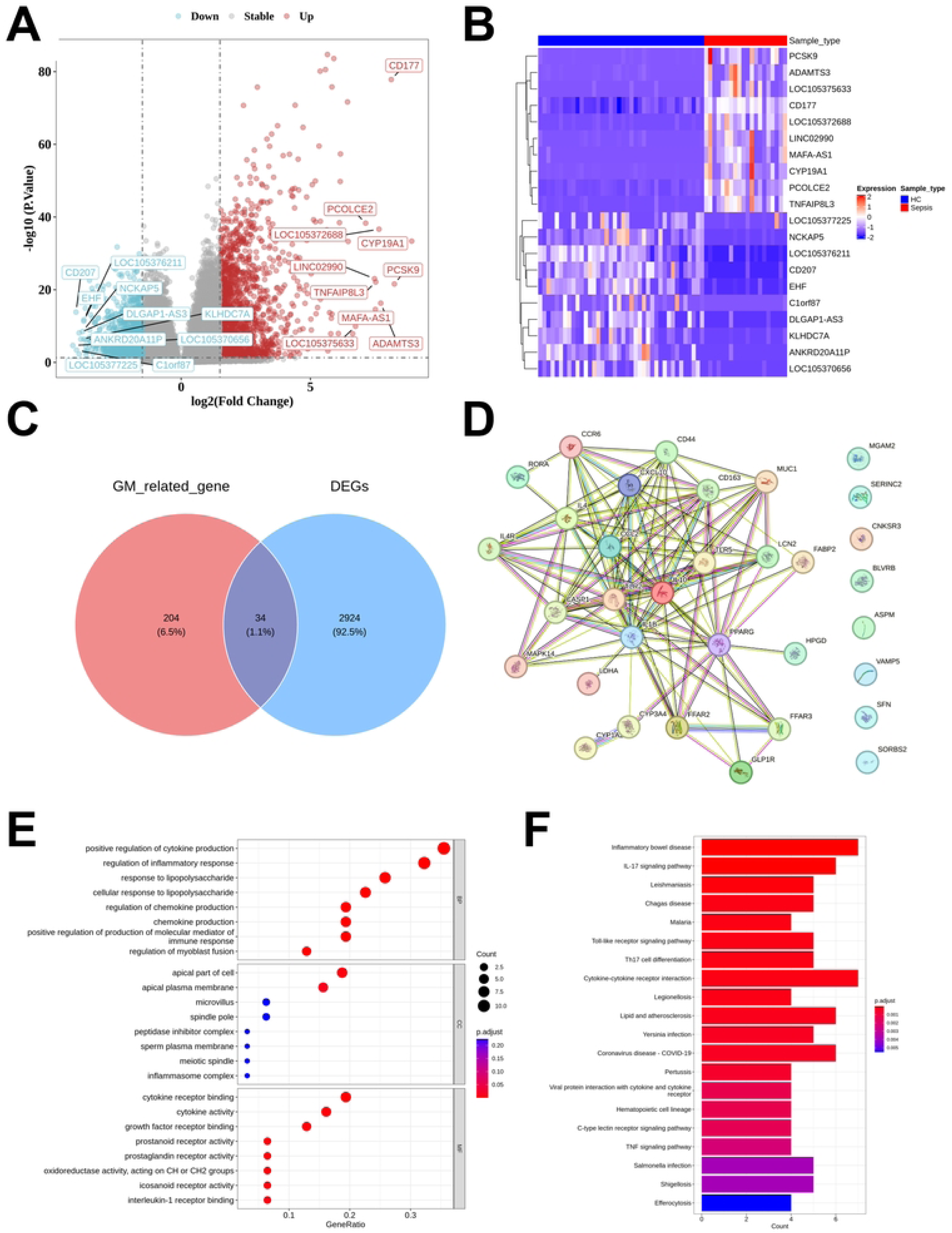
Identification of gut microbiota-related DEGs between sepsis and healthy control, followed by PPI, GO and KEGG analysis. (A) DEGs shown on a volcano plot in GSE154981. (B) Heatmap demonstration of Top 10 genes identified via differential expression analysis. (C) Venn diagram showed the intersected genes of gut microbiota-related genes and DEGs. (D) PPI network of 34 gut microbiota-related DEGs. (E) GO functional enrichment analysis of 34 gut microbiota-related DEGs. (F) KEGG pathway enrichment analysis of 34 gut microbiota-related DEGs.

GO analysis revealed significant enrichment in biological processes such as positive regulation of cytokine production, regulation of inflammatory response, response to lipopolysaccharide, and chemokine production. Additionally, enrichment was also observed in specific cellular components including the apical part of cell, apical plasma membrane, and inflammasome complex. Furthermore, enrichment was noted in molecular functions such as cytokine receptor binding, cytokine activity, and prostaglandin receptor activity (Fig. 2E). The KEGG pathway enrichment analysis revealed that the candidate genes were mainly involved in pathways such as inflammatory bowel disease, IL-17 signaling pathway, and cytokine-cytokine receptor interaction (Fig. 2F).

### Construction and Validation of the Prognostic Model (GMGscore)

Using GSE65682 as the training set, univariate Cox analysis identified 10 genes significantly associated with prognosis (Fig. 3A). Subsequently, LASSO regression analysis (λ = 0.01) was performed to screen 6 genes (CYP1A2, FFAR2, IL4R, MUC1, RORA, and ASPM) from the 10 candidate genes that were significantly correlated with 28-day mortality in sepsis patients, and the corresponding prognostic model was designated as GMGscore (Figs. 3B–C). The genes included in GMGscore and their respective coefficients are provided in Table S3.

**Fig. 3.**
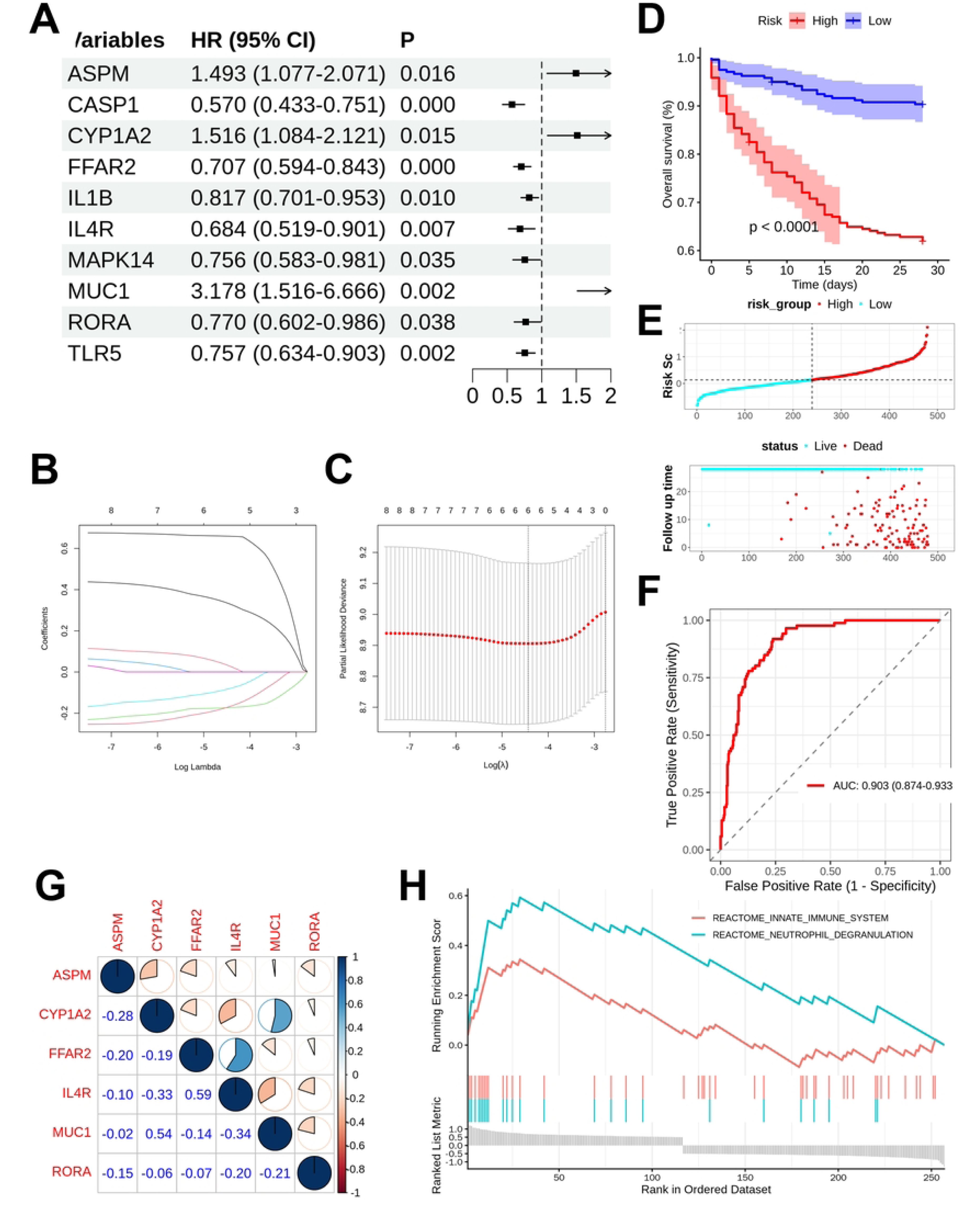
Construction and validation of the GMGscore. (A) Univariate cox regression for identifying the prognosis-related genes in the gut microbiota-related DEGs. (B) LASSO coefficient path plot. (C) Cross-validation curve of LASSO regression analysis. (D) Survival analysis showed a significantly worse prognosis in the high-GMGscore group. (E) The distribution of GMGscore and outcome status. (F) The ROC curve of the training cohort demonstrated AUC value of 0.903. (G) Spearman correlation among the six prognosis genes. (H) GSEA results associated with GMGscore.

Patients in the dataset were divided into high-risk and low-risk groups based on the median GMGscore. Kaplan-Meier (KM) analysis demonstrated that sepsis patients in the high-risk group had significantly poorer prognosis (P < 0.001) (Fig. 3D). The GMGscore risk score-death event distribution plots (Figs. 3E–F) and heatmap (Fig. S2) further confirmed that higher GMGscore was associated with worse prognosis. Additionally, subgroup survival analyses were conducted to verify the generalizability of GMGscore. The results showed that GMGscore exhibited robust prognostic discrimination ability across subgroups of male (Fig. S3A), female (Fig. S3B), elderly (Fig. S3C), non-elderly (Fig. S3D), diabetic (Fig. S3E), and non-diabetic (Fig. S3F) sepsis patients. ROC analysis indicated that GMGscore had excellent performance in prognostic prediction in the training set (AUC = 0.903, 95% confidence interval [CI]: 0.874–0.933) (Fig. 3G). Validation using the GSE95233 dataset (Fig. S4) confirmed the strong generalization ability of GMGscore (AUC = 0.901, 95% CI: 0.813–0.990). Spearman correlation analysis was performed to evaluate the expression correlations of the 6 key prognostic genes in sepsis samples (Fig. 3H). Significant positive correlations were observed between FFAR2 and IL4R (R = 0.59), and between MUC1 and CYP1A2 (R = 0.54). In contrast, IL4R showed significant negative correlations with MUC1 (R = −0.34) and CYP1A2 (R = −0.33).

To explore the biological significance associated with GMGscore, GSEA was conducted. The results showed that the “innate immune system” (normalized enrichment score [NES] = 2.187, P = 0.001) and “neutrophil degranulation” (NES = 3.089, P < 0.001) were significantly upregulated in samples with high GMGscore. Furthermore, immune infiltration analysis revealed that sepsis patients in the high-GMGscore group had fewer neutrophils (P < 0.001) (Fig. S5). Spearman correlation test also confirmed a significant negative correlation between GMGscore and neutrophil levels in sepsis patients (R = −0.304, P < 0.001) (Fig. S6). In addition, compared with the low-GMGscore group, the high-GMGscore group had a reduction in activated mast cells and an increase in resting mast cells (P < 0.001).

### Association Between GMGscore and Clinical Variables, and Assessment of the Nomogram

GMGscore was compared across different subgroups to assess its association with clinical variables. The results showed no significant associations between GMGscore and age (Figs. 4A, S7A), diabetes status (Fig. 4B), or gender (Figs. 4C, S7B–C). However, GMGscore was significantly higher in sepsis patients than in healthy controls across all datasets (Figs. 4D, S7D–F).

**Fig. 4.**
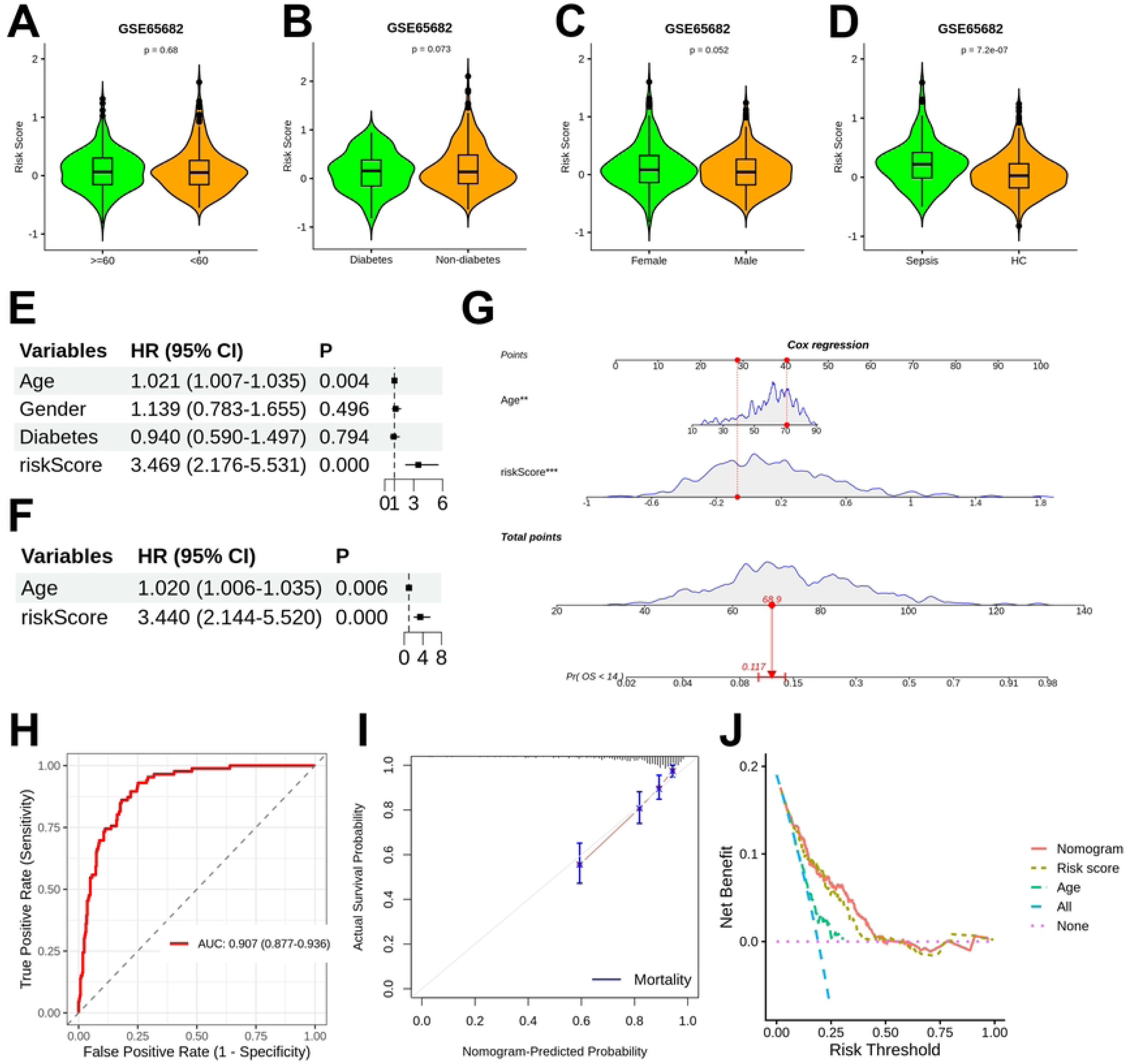
Association between GMGscore and linical variables, and the construction of a nomogram. (A) Violin plot showing that no significant difference was found between the GMGscores of sepsis patients younger than 60 years and those aged 60 years or older. (B) Violin plot illustrating no significant difference in GMGscores between diabetic and non-diabetic sepsis patients. (C) Violin plot demonstrating no significant difference in GMGscores between male and female sepsis patients. (D) Violin plot illustrating significant difference in GMGscores between healthy individuals and sepsis patients. (E) Univariate Cox regression for identifying the prognosis-related variables. (F) Multivariate Cox regression results of age and GMGscore. (G) A nomogram was developed via integrating clinical data and GMGscores. (H) ROC analysis for the nomogram showed the AUC value of 0.907. (I) Calibration curve for the nomogram. (J) DCA result demonstrated superior performance compared to single indicators.

Univariate and multivariate Cox regression analyses were performed by integrating clinical variables (Figs. 4E–F). Based on these results, a nomogram was constructed using GMGscore and age (HR = 1.021, P = 0.002) as predictive parameters (Fig. 4G). ROC analysis showed that the nomogram had an AUC of 0.907 (95% CI: 0.877–0.936) for prognostic prediction (Fig. 4H). Calibration curves demonstrated the excellent prognostic performance of the nomogram (Fig. 4I). DCA revealed that the nomogram provided greater clinical benefits than GMGscore or age alone (Fig. 4J). Collectively, these results suggest that GMGscore has the potential to serve as a reliable clinical tool for sepsis prognosis prediction.

### Screening of Key Targets and Molecular Docking with Potential Therapeutic Metabolites

We visualized the regulatory relationships between prognostic genes and gut microbiota/metabolites (Fig 5A). In the visualization, red lines represent activation, blue lines represent inhibition, dashed lines represent correlation, and solid lines represent causal association. Notably, only urolithin A exerted an inhibitory effect on ASPM.

**Fig. 5.**
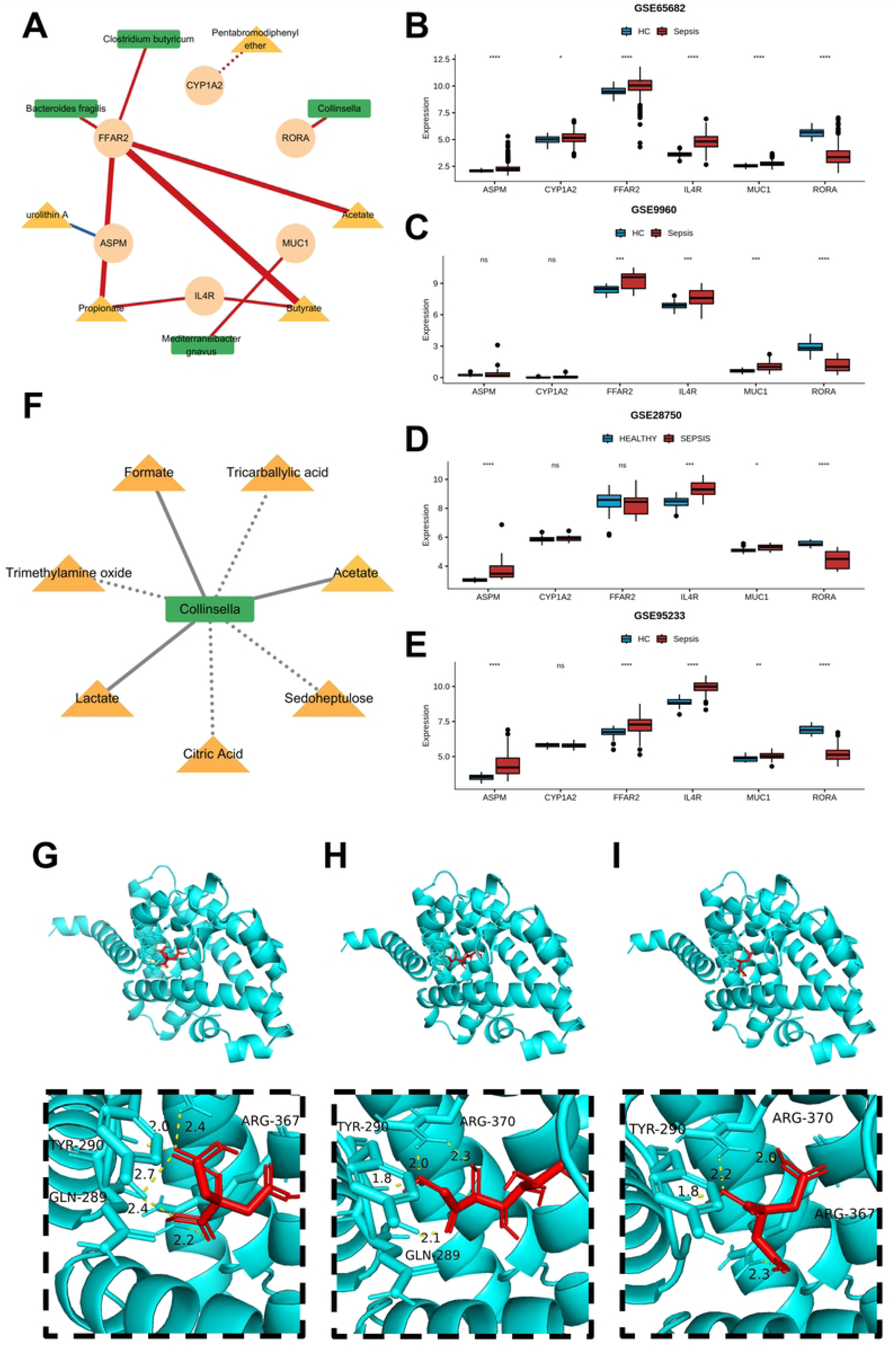
Screening of potential target for sepsis and promising metabolites for the treatment. (A) Network for demonstrating the relationship among six prognosis genes and microbiota/metabolites. (B) Boxplot showing the expression level of six prognosis genes in GSE65682. (C) Boxplot showing the expression level of six prognosis genes in GSE9960. (D) Boxplot showing the expression level of six prognosis genes in GSE28750. (E) Boxplot showing the expression level of six prognosis genes in GSE95233. (G) Molecular docking result of RORA and Citric acid. (H) Molecular docking result of RORA and Sedoheptulose. (I) Molecular docking result of RORA and Tricarballylic acid.

We further verified the consistency of prognostic gene expression across four datasets: GSE65682, GSE95233, GSE28750, and GSE9960. The results showed that IL4R and MUC1 were consistently highly expressed in sepsis patients, while Retinoic Acid Receptor-Related Orphan Receptor Alpha (RORA) was consistently lowly expressed (P<0.001, Fig 5B-D). Additionally, RORA exhibited the highest AUC value for sepsis diagnosis across all four datasets (Fig S8A-D). In the interaction network, we observed that Collinsella could activate RORA, which aligns with the direction of improved sepsis prognosis (Fig 5A). Therefore, we consider RORA a potential target that can be intervened via the gut microbiota.

Collinsella was found to have correlational relationships with 7 metabolites: Acetate, Citric acid, Formate, Lactate, Sedoheptulose, Tricarballylic acid, and Trimethylamine oxide (Fig 5E). Molecular docking results (Table S4) revealed that three of these metabolites formed relatively stable bindings with RORA protein: Citric acid (binding energy = −5.5 kcal/mol), Sedoheptulose (binding energy = −5.2 kcal/mol), Tricarballylic acid (binding energy = −5.6 kcal/mol). Notably, the predicted binding sites of all three metabolites included the tyrosine residue at position 290 (Fig 5F-H).

### Single-Cell RNA-seq Data Analysis

Using the UMAP method, all cells were clustered into 25 clusters and annotated into 7 cell types: NK cells, T cells, B cells, neutrophils, myeloid cells, platelets, and erythroid cells (Fig 6B). The differential expression of marker genes across different cell types confirmed the reliability of cell clustering (Fig 6D).

**Fig. 6.**
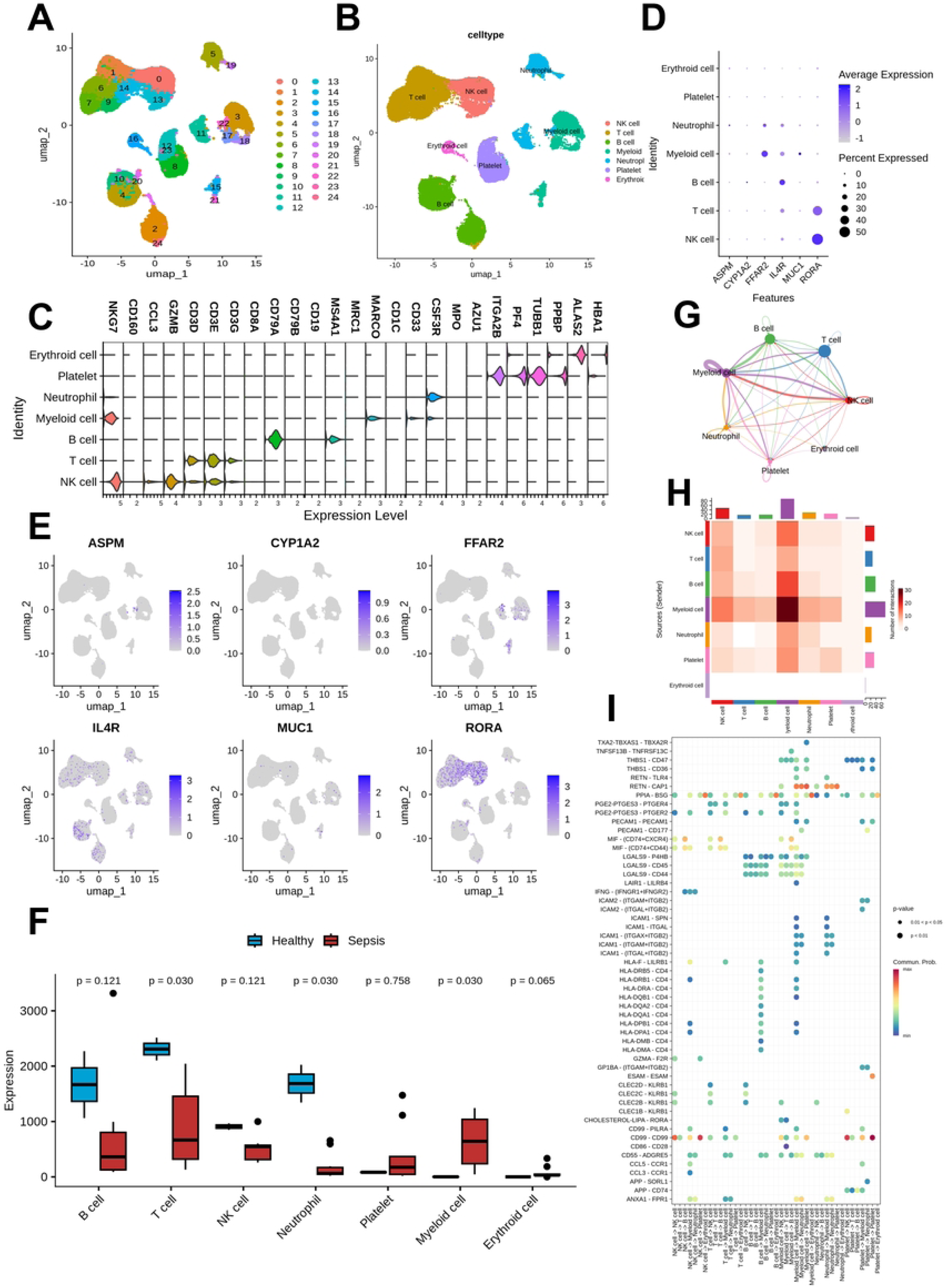
Single-cell RNA sequencing data analysis. (A) A total of 25 clusters were identified using UMAP clustering. (B) Cell types of the dataset. (C) Marker genes of different cell clusters. (D) Dot plot showing the expression levels of the six prognosis genes in different cell types. (E) UMAP plot demonstrating the cell types distribution of the six prognosis genes. (F) Comparison for the cell counts of different cell types between sepsis and healthy control. (G) Network plot result of cell communication analysis. (H) Heatmap result of cell communication analysis. (I) Bubble plot showing the ligand-receptor pair of cell-cell interaction.

We then mapped the distribution of prognostic genes in these cells (Fig 6D-E), and the analysis showed that RORA was mainly expressed in T cells and NK cells. By comparing the counts of various cell types between healthy samples and sepsis samples, we found that T cells, neutrophils and myeloid cells were significantly reduced in sepsis samples; thus, we focused on T cells in subsequent analyses.

CellChat was used to visualize the intercellular communication network in sepsis samples (Fig 6G-H), which showed that myeloid cells had a higher communication probability with other cell types. Additionally, the ligand-receptor pairs mediating intercellular communication in sepsis were identified and presented (Fig. 6I).

To further explore T cells, we subclassified the focused T cells into subsets (Fig 7A-B). The results indicated that RORA was primarily distributed in effector T cells (Fig. 7C). Immune infiltration analysis (Fig. 7D) revealed that samples with high RORA expression had higher immune infiltration levels of CD8+ T cells and activated NK cells, while the infiltration levels of naive CD4+ T cells and resting NK cells were lower. Spearman correlation analysis (Fig 7E, Fig S9A-B) demonstrated that RORA was significantly positively correlated with the levels of CD8+ T cells (R = 0.419, P<0.001) and activated NK cells (R = 0.352, P<0.001).

**Fig. 7.**
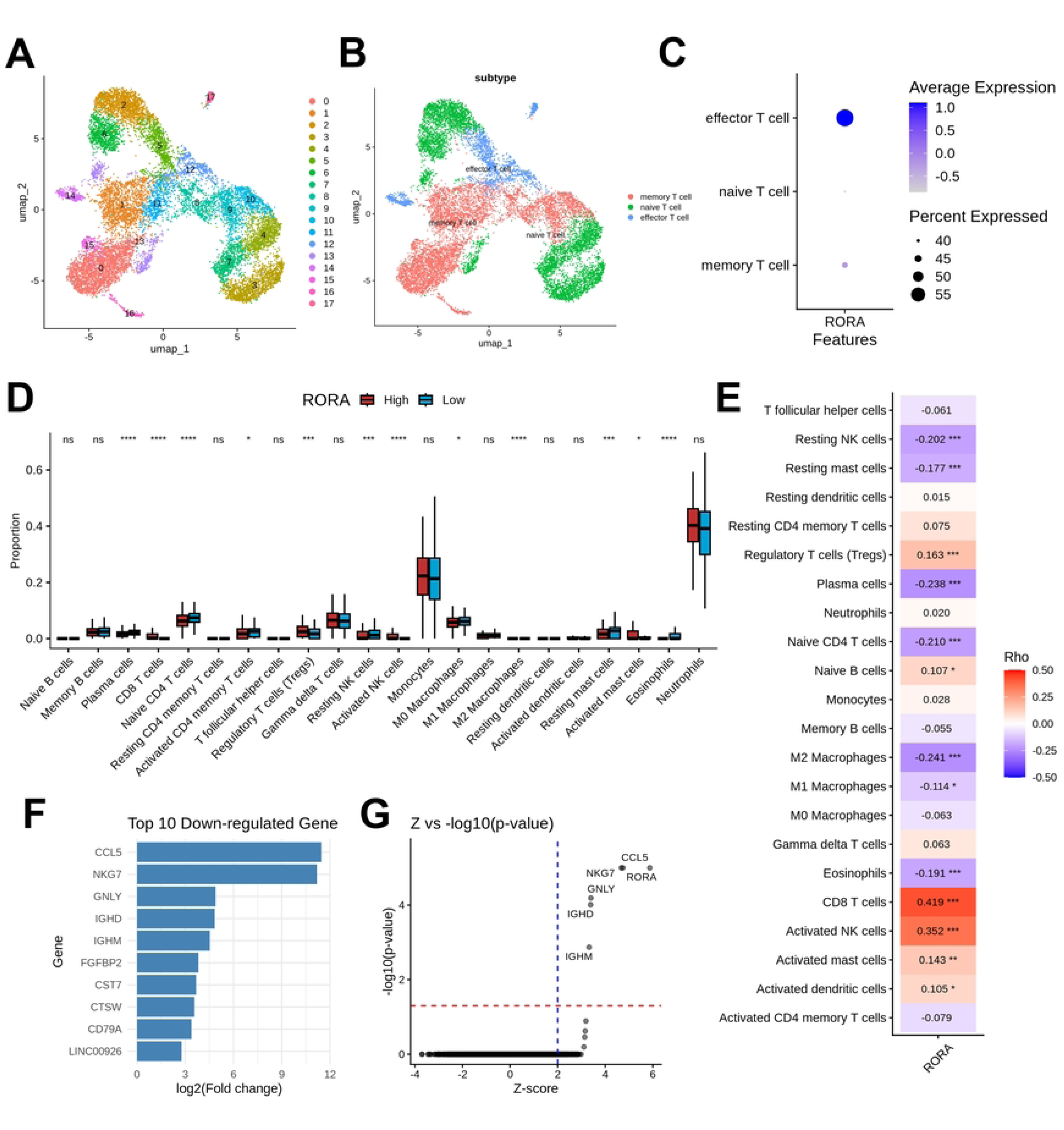
The role of RORA in sepsis was inferred through multiple analytical methods. (A) A total of 18 clusters were identified from T cell subset using UMAP clustering. (B) T cell subtypes of the subset. (C) Dot plot showing the RORA expression level among different T cell subtypes. (D) Immune infiltration analysis associated with RORA expression. (E) Spearman correlation between RORA and 22 types of immune cells. (F) Bar plot showing the top 10 down-regulated genes after virtual KO of RORA. (G) Volcano plot showing significantly dysregulated genes following virtual KO of RORA.

We also conducted virtual knockout analysis (Fig 7F-G). After virtual knockout of RORA, the expression levels of genes such as CCL5, NKG7, and GNLY were significantly downregulated. This suggests that RORA may influence the immune balance in sepsis by regulating the function or recruitment of effector T cells and NK cells.

## Discussion

Risk stratification of sepsis serves as a critical basis for early targeted intervention, and gut microbiota-associated biomarkers offer a more comprehensive perspective for prognostic assessment in critical care medicine (15). Against the backdrop of the big data era, multi-omics data has provided robust support for the prediction of sepsis prognosis (27).

Through integrated multi-omics analysis, this study constructed, for the first time, a sepsis prognostic model (GMGscore) based on gut microbiota-related genes. The model demonstrated excellent performance in both the training set and validation set, with AUC values reaching 0.903 and 0.901, respectively. It is expected to be applied in clinical practice to identify high-risk sepsis patients, thereby enabling early targeted treatment. For comparison, the 28-day mortality prediction model based on autophagy-related genes developed by Chen et al. had an AUC of 0.700 (28), while their HLA-related classifier showed an AUC range of 0.691–0.752 (29). Jiang et al. established a prognostic model using an inflammatory response-related gene signature, which achieved an AUC of 0.866 (30), and the prognostic model constructed by Jiang et al. had an AUC range from 0.707 to 0.856 (31). These results further underscore the significance of gut microbiota in sepsis prognosis.

We explored the correlations of GMGscore with clinical features, immune infiltration, and pathway enrichment. In the analysis of clinical correlations, GMGscore showed no association with age, gender, or the presence of diabetes, indicating that GMGscore is a relatively independent prognostic indicator. Consistently, GMGscore was elevated in sepsis patients. Immune infiltration analysis revealed that the number of neutrophils decreased with increasing GMGscore (R = −0.304, P < 0.001), and scRNA seq analysis showed a similar result (P = 0.030). A reduction in neutrophil count may imply impaired anti-infective capacity, thereby contributing to the progression of sepsis (32). GSEA pathway enrichment analysis showed upregulation of pathways such as neutrophil degranulation and the innate immune system. The activation of neutrophil degranulation and the innate immune system are hallmark upregulated pathways in early sepsis, which may indicate excessive activation of these biological processes in high-risk sepsis patients (33). Excessive activation of neutrophil degranulation, while effective in killing pathogens, can also damage the normal tissues and organs of the organism (34). Notably, neutrophil dysfunction is a core pathological event in sepsis (35). These findings suggest that the high-risk status defined by GMGscore may be mediated through changes in neutrophil quantity and function.

The genus Collinsella is typically described as a strictly anaerobic pathogenic bacterium that produces lactic acid rather than butyrate or other short-chain fatty acids (36). It is closely associated with non-alcoholic steatohepatitis and cholesterol metabolism and is considered a pro-inflammatory pathogenic genus (37, 38). However, the results of our study suggested that Collinsella may improve sepsis prognosis by activating RORA through its metabolites.

This study also identified RORA as a potential target amenable to gut microbiota-based intervention. RORA is a transcription factor belonging to the nuclear receptor superfamily, and it plays roles in multiple processes including neural function, cell development, immune regulation, metabolism, and circadian rhythm (39). Previous studies have confirmed that RORA is involved in inflammatory diseases by regulating the circadian clock and immune cell activation (40). Upregulation of RORA can prevent inflammation and inhibit the expression of adhesion-related proteins, including intercellular adhesion molecule 1 (ICAM-1) and vascular cell adhesion molecule 1 (VCAM-1) in human umbilical vein endothelial cells (HUVECs) (41).

RORA expression is downregulated in sepsis patients and is regarded as a key regulator (42, 43). Previous studies have shown that RORA exhibits high diagnostic performance for sepsis (44), which is consistent with the findings of our study. Experiments in mice have confirmed that RORA is a key factor in initiating the innate immune response against inflammation and exerts a protective role during the inflammatory process (45). The present study found that RORA is lowly expressed in sepsis patients and is significantly associated with poor prognosis. Besides, RORA is mainly enriched in effector T cells and NK cells. Additionally, gut microbiota metabolites including citric acid, sedoheptulose, and tricarballylic acid may bind stably to RORA, suggesting that gut microbiota may regulate the immune response in sepsis patients through the metabolite-RORA interaction.

In our study, the significant positive correlation between RORA and CD8+ T cells/NK cells highlights its positive role in regulating immune cell function. Furthermore, results from virtual knockdown of RORA showed that low RORA expression downregulates the expression of cytotoxic genes such as CCL5, NKG7, and GNLY. This may restrict the recruitment and functional exertion of T cells and NK cells, thereby mediating immune suppression (46, 47). In animal experiments, upregulation of RORA expression using sevoflurane alleviated lipopolysaccharide-induced endothelial cell damage (48). Targeting RORA to restore circadian rhythm may represent an innovative therapeutic approach to alleviate immune dysfunction and improve patient prognosis (49).

This study has certain limitations. First, it relies on public datasets, and the samples are mainly derived from European populations. The lack of validation in Asian populations may affect the racial applicability of the model. Second, the sample size of scRNA-seq data is relatively small, so the cellular localization results of RORA need to be verified with an expanded sample size. Third, the study is limited to bioinformatics analysis; cell and animal experiments are required to validate the regulatory role of RORA expression and activity on the body’s immune system, as well as its impact on sepsis-related inflammatory factors. Finally, this study did not include clinical samples for protein-level validation. Future multi-center clinical studies are needed to verify the clinical utility of GMGscore and RORA-targeted interventions.

## Conclusions

Through integrated analysis of transcriptome and single-cell sequencing data, this study constructed a sepsis prognostic model (GMGscore) based on 6 gut microbiota-related genes. This model exhibits excellent prognostic discrimination ability and holds potential clinical application value. Meanwhile, RORA was identified as a key target for gut microbiota-based intervention. It may influence the immune balance in sepsis by regulating the functions of effector T cells and NK cells. These findings not only deepen the understanding of the mechanism by which gut microbiota contributes to sepsis progression but also provide a novel perspective for the precise prognostic assessment and targeted treatment of sepsis. Multi-center clinical validation and functional experiments are required to further promote the clinical translational application of GMGscore and RORA.

## Abbreviations

AUC: area under the curve
BP: Biological Process
CC: Cellular Component
DCA: decision curve analysis
DEG: Differentially expressed gene
GEO: Gene Expression Omnibus
GMGscore: gut microbiota related genes score
GO: Gene Ontology
GRN: gene regulatory network
GSEA: Gene Set Enrichment Analysis
HUVEC: human umbilical vein endothelial cell
ICAM-1: intercellular adhesion molecule 1
ICU: intensive care units
KEGG: Kyoto Encyclopedia of Genes and Genomes
KM: Kaplan-Meier
KO: knockout
MF: Molecular Function
PPI: Protein-Protein Interaction
RORA: Retinoic Acid Receptor-Related Orphan Receptor Alpha
scRNA-seq: Single-Cell RNA Sequencing
UMAP: Uniform Manifold Approximation and Projection
UMI: unique molecular identifier
VCAM-1: vascular cell adhesion molecule 1

## Ethics and approval

The research was conducted in full compliance with the guidelines outlined in the Declaration of Helsinki. This study was approved by the Medical Ethics Committee of Tongde Hospital of Zhejiang Province.

## CRediT authorship contribution statement

Investigation: Fangqiong Li; Writing-original manuscript: Fangqiong Li, Minrong Xu; Data extraction and statistical analysis: Fangqiong Li, Huiqin Xiao; Writing-reviewing and editing: Fangqiong Li, Ping Hu, and Wei Zhang; Project administration: Wei Zhang; Conceptualization and supervision: Wei Zhang. All authors have read and agreed to the published version of the manuscript.

## Data availability

The datasets used for all analyses in this research are publicly available on GEO website (https://www.ncbi.nlm.nih.gov/geo/).

## Acknowledgments

We appreciate the GEO databases for their platforms and contributors for uploading their significant datasets.

